# A Simple Mathematical Model for Estimating the Inflection Points of COVID-19 Outbreaks

**DOI:** 10.1101/2020.03.25.20043893

**Authors:** Zhanshan Sam Ma

## Abstract

**Background:** Exponential-like infection growths leading to peaks (which could be the inflection points or turning points) are usually the hallmarks of infectious disease outbreaks including coronaviruses. To predict the inflection points, *i.e*., inflection time (*T*_max_) & maximal infection number (*I*_max_) of the novel coronavirus (COVID-19), we adopted a trial and error strategy and explored a series of approaches from simple *logistic modeling* (that has an asymptomatic line) to sophisticated *tipping point* detection techniques for detecting phase transitions but failed to obtain satisfactory results.

**Method:** Inspired by its success in diversity-time relationship (DTR), we apply the PLEC (power law with exponential cutoff) model for detecting the inflection points of COVID-19 outbreaks. The model was previously used to extend the classic species-time relationship (STR) for general DTR (Ma 2018), and it has two “secondary” parameters (computed from its 3 parameters including power law scaling parameter *w*, taper-off parameter *d* to overwhelm virtually exponential growth ultimately, and a parameter *c* related to initial infections): one that was originally used for estimating the potential or ‘dark’ biodiversity is proposed to estimate the *maximal infection number* (*I*_*max*_) and another is proposed to determine the corresponding *inflection time point* (*T*_*max*_).

**Results:** We successfully estimated the inflection points [*I*_max_, *T*_max_] for most provinces (≈85%) in China with error rates <5% in both *I*_*max*_ and *T*_*max*_. We also discussed the constraints and limitations of the proposed approach, including (*i*) sensitive to disruptive jumps, (*ii*) requiring sufficiently long datasets, and (*iii*) limited to unimodal outbreaks.

## Introduction

In spite of the obviously extreme importance and extensive research efforts, predicting the inflection points, formally known as the tipping points or turning points in much of the existing literature, turned out to be notoriously difficult. The enormous complexity associated with tipping points and critical transitions make accurate predictive modeling nearly impossible. Existing theories suggest that early warning signals (EWS) may be developed based on the phenomenon that recovery rates from small disturbances should tend to zero when approaching a tipping point (Scheffer et al. 2009, 2015), and some laboratory experiments and field survey have demonstrated the so-called “critical slow down” (CSD) phenomenon (*e.g*., Veraart et al. 2012, Dakos *et al*. 2015).

In theory, it has been suggested that even without fully mechanistic understanding of the underlying critical transitions, it is still possible to infer generic features or early warning signals (EWS) of approaching a tipping point from time series or spatial pattern data. Theory proposes that at bifurcation points (*i.e*., tipping points), the stability of an equilibrium changes and the dominant real eigenvalue becomes zero. Consequently, the recovery rate from disturbance should go zero when approaching such bifurcation, or the return time (measure of resilience) should be infinity (Veraart *et al*. 2012). In practice, the recovery rate from disturbance may be an indicator of the distance to a tipping point, and its manifestation, the CSD offer important early EWS for approaching a tipping point.

Investigating the inflection points of infectious disease outbreaks including the ongoing coronaviruses pandemic falls under the domain of tipping points and critical transitions (Veraart et al. 2012, Dakos *et al*. 2015, Scheffer et al. 2009, 2015). However, in a pre-experiment analysis of the existing datasets of COVID-19 infections, we failed to detect any tipping points (inflection points). We conjecture that the failure may have to do with the insufficiently long duration of data collections. We then also tried some simple epidemiological modeling approaches such as simple logistic model (*e.g*., Ma *et al*. 2009) but still failed to achieve satisfactory results.

Inspired by the exponential-like growth of infections leading to peaks, which seems to be a hallmark of many infectious disease outbreaks including COVID-19 and SARS (Rivers *et al*. 2019, Li *et al*. 2020, Thompson 2020, Ma 2020), we try the PLEC (power law with exponential cutoff) model for detecting the inflection points in the time series data of COVID-19. The PLEC model (function) starts with power function increase (near exponential growth usually) and it has an exponential decay term that acts as taper-off parameter to eventually overwhelm the power law behavior at very large value of independent variable. The PLEC model was previously used to describe the classic species-area relationship (SAR) in biogeography (Plotkin *et al*. 2000, Ulrich & Buszko 2003), and later used for extending the classic SAR and STR (species-time relationship) to general diversity-area relationship (DAR) and diversity-time relationship (DTR) (Ma 2018a, 2018b, 2019). Furthermore, Ma (2018a, 2018b, 2019) derived the formula to estimate the maximal accrual diversity (MAD) in space (area), time, and/or spatiotemporal settings, which may also be used to estimate potential diversity (also known as “dark” diversity). The potential or dark diversity refers to the biodiversity that take into accounts the contribution of species that are absent locally but potentially present in regional species pool. This situation of potential species migration from regional (metacommunity) to local community is not unlike the COVID-19 infections via dispersal (migration) such as travelers of virus-infected individuals. We postulate that the PLEC model can be useful for modeling the infection-time relationship (ITR)—the relationship between the cumulative number of infections and time points (*e.g*., day 1, 2,……,*N*). If the PLEC model successfully fits to the ITR datasets, we further postulate that the maximal accrual diversity or potential diversity and corresponding time point may be translated into an approach for estimating the *maximal infection number* (*I*_max_) of an outbreak and the corresponding time point (when *I*_max_ is reached) as estimate of inflection time point (*T*_max_). Together the pair of {*I*_max_, *T*_max_} is defined as the inflection point of a disease outbreak in this study. We further devised schemes to test the performance of the PLEC model in predicting (estimating) the inflection points of COVID-19 outbreaks.

## Material and Methods

### The PLEC model (Power Law with Exponential Cutoff)

A hallmark of coronaviruses outbreaks seems to be exponential or near exponential growth of infections, and the peak time and amplitude or the outbreak inflection points of the virus are of obvious importance both theoretically and practically. We address this problem with a trial-and-error strategy. We first conjectured that simple logistic growth model could be a possible solution since its asymptotic limit (*K*) might be used to approximate the inflection points (Ma *et al*. 2009). We also tried sophisticated mathematical approaches for detecting tipping points (Dai *et al*. 2012, 2013, Sundstrom *et al*. 2017). However, the applications of both logistic growth model and tipping point approaches failed to generate satisfactory results.

In the present report, we adopt the PLEC (power law with exponential cutoff) model for estimating the inflection points of coronavirus infections. The PLEC model is of the following form:

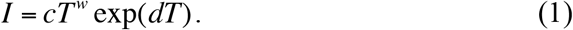

The model was initially used by Plotkin *et al* (2000) and Ulrich & Buszko (2003) for modeling the classic species-area relationship (SAR). Ma (2018a, 2018b, 2019) introduced it for more general diversity-area relationship (DAR), diversity-time relationship (DTR) and diversity-time-area (DTAR) relationship. Another extension to the model was the derivation of two parameters by Ma (2018a, 2018b, 2019): the maximal accrual diversity and corresponding time or space points. The maximal accrual diversity can be used to estimate the so-termed potential diversity (also known as “dark” diversity), which takes into accounts of the species that are absent locally (in local communities) but present regionally (in metacommunities) (Ma 2019, Li & Ma 2019).

In the present report, we use the PLEC (power law with exponential cutoff) model (eqn. 1) for modeling the growth of COVID-19 infections. With eqn. (1), *T* is the time or days (starting from a specified date), and *I* is the number of cumulative infections, *c, d*, and *w* are three parameters to be estimated from the observed data. Ma (2018a) derived the maximum of eqn. (1) by solving the following equation:

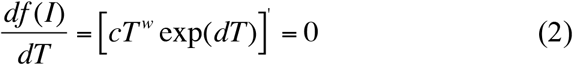

which leads to:

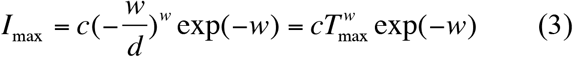

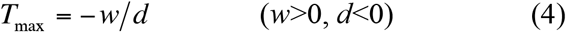

Theoretically, eqns. (3) & (4) can be utilized to estimate the *maximal infection number* (*I*_max_) and corresponding *inflection time* point (*T*_max_) at which *I*_max_ occurs.

Parameter *d* is of particular interests: *exp*(*dT*) in eqn. (1) is the exponential decay term that acts as taper-off parameter to eventually overwhelm the power law behavior (exponential or near exponential growth) at very large value of *T*. For eqn. (*1*) to achieve maximum of realistic biomedical meaning (*T*_max_>0) or (*w*>0 & *d*<0) are necessary conditions.

The biological interpretation of parameter *w* & *c* can best be observed by assuming *d*→0. When *d*=0, the PLEC defaults to classic power-law model,

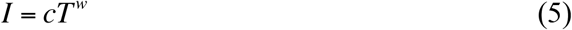

or its log-transformed linear model:

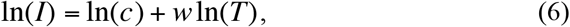

in which *w* is the slope of the log-linear model or the derivative of *I*(*T*), *i.e*., the change (growth) rate of infections over time, and *c* is the initial infection number. Since the taper-off effects of parameter *d* is usually rather weak before the infection peaks, it is reasonable to consider *w* as an approximation of the infection growth rate, and *c* as an approximation of the initial infection number.

A simple method to statistically fit the PLEC model [eqn. (1)] is to transform it into a log-linear equation as follows:

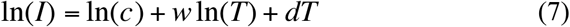

For eqn. (7), simple linear regression can be used to fit the model and hence obtain the PLEC parameters. Nevertheless, the simple linear regression may not provide satisfactory fitting to the original PLEC model [eqn. (1)], especially when the number of data points for fitting the model is small or moderate. For this reason, we use the nonlinear optimization method to fit the PLEC model [(eqn.1)] directly, rather than fitting the log-linear transformed equation [eqn. (7)] indirectly. We used nonlinear optimization method, specifically the R function “nlsLM” from R package “minpack.lm” (https://www.rdocumentation.org/packages/minpack.lm/versions/1.2-1/topics/nlsLM) (Bates & Watts 1988). Since only *T*_max_>0 (negative time is meaningless to count infections) is a necessary condition for the PLEC model to be biometrically sound, we add a constraint *d*<0 for the non-linear fitting of PLEC model.

It is particularly worthy of noticing that the dependent variable (*I*) of PLEC, almost unavoidably, declines after the inflection point (maximum) due to the taper-off parameter (*d*). Nevertheless, this should not be an issue in the case of this study, since our purpose is to detect the inflection point (*T*_max_) as well as the corresponding maximum (*I*_max_). In other words, the declining piece of PLEC curve after the inflection point is “irrelevant” for our purpose. Of course, this also implies that our model is only applicable to unimodal (single peak) outbreak.

### The COVID-19 infection datasets and criteria for evaluating the PLEC model

We collected the daily infections datasets of COVID-19 in 34 Chinese provinces and the world from the public domain. In addition, we collected the datasets of 17 cities of Hubei province of China. The infection data between Jan 19^th^ and Feb 29^th^ were collected for building the predictive PLEC model, and the infection data at two time points (March 6^th^ and 12^th^) as the test data points to evaluate the performance of the PLEC model in determining the inflection points. The public websites used to get the data include: https://news.qq.com/zt2020/page/feiyan.htm#/ and https://news.ifeng.com/c/special/7tPlDSzDgVk.

We test the performance of the proposed PLEC model with two schemes. First, we evaluate the “congruency” of *T*_max_ (inflection time point) with the reality by comparing the *actually observed* infection number at the date of *T*_max_ with the estimated *I*_max_. To determine the congruence, we calculate an interval [*I*_max_−*I*_max_x0.05; *I*_max_+*I*_max_x0.05] for each PLEC model. If the actually observed infection numbers falls within the interval, the estimation of *T*_max_ is considered as valid. It is noted that this is *not* a classic *confidence interval*, which requires the calculation of standard deviation of samples and cannot be obtained in the case of COVID-19 infections. Instead, this “homegrown” interval we computed is simply a range of *I*_max_ with 5% fluctuation, which we believe is a reasonable scheme for estimating the validity of the *T*_max_ estimations.

Second, to evaluate the validity of *I*_max_ estimations, we used the observed infection number on the day of *T*_max_, March 6^th^ and March 12^th^, respectively, as reference values to evaluate the performance of the PLEC model, using the following formula to calculate the error rate:

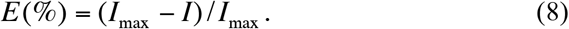

Alternatively, the precision *P*(%) of the PLEC model is simply computed as the complement of *E*(%), *i.e*., *P*(*%*)*=1–E*(*%*).

## Results

We fitted the PLEC model with the nonlinear optimization method (*i.e*., the R-function ‘nlsLM’ introduced in the previous “The Model” section) to each of the 17 cities in Hubei province, each of the 34 provinces of China, as well as the whole China and worldwide infections datasets. The results were displayed in Table 1 (the PLEC model-fitting results) and Table 2 (test of the model performance). Since the model mostly failed to make meaningful predictions of the infections outside China, we postpone the worldwide results to the “Discussion” section and here we focus on the results fitted to the datasets in China.

**Table 1.** The ITR-PLEC (Infection Time Relationship—Power Law with Exponential Cutoff) model, fitted with nonlinear optimization for daily cumulative COVID-19 infections*

| Data | Country /Province | $z$ | $z(p)$ | $d$ | $d(p)$ | $c$ | $c(p)$ | $R^2$ | $T_{max}$ | $I_{max}$ |
| --- | --- | --- | --- | --- | --- | --- | --- | --- | --- | --- |
| Hubei | Wuhan | 9.006 | 0.000 | -0.188 | 0.000 | 0.000 | 0.638 | 0.986 | 48 | 49193.4 |
|  | Xiaogan | 2.856 | 0.000 | -0.091 | 0.000 | 3.232 | 0.001 | 0.993 | 32 | 3534.6 |
|  | Huanggang | 3.007 | 0.000 | -0.094 | 0.000 | 1.806 | 0.000 | 0.994 | 32 | 2979.4 |
|  | Jingzhou | 3.288 | 0.000 | -0.102 | 0.000 | 0.468 | 0.005 | 0.993 | 32 | 1607.0 |
|  | Ezhou | 3.544 | 0.000 | -0.110 | 0.000 | 0.221 | 0.024 | 0.992 | 32 | 1413.2 |
|  | Suizhou | 2.340 | 0.000 | -0.081 | 0.000 | 5.357 | 0.000 | 0.992 | 29 | 1336.7 |
|  | Xiangyang | 1.876 | 0.000 | -0.070 | 0.000 | 16.522 | 0.000 | 0.992 | 27 | 1210.0 |
|  | Huangshi | 2.268 | 0.000 | -0.081 | 0.000 | 5.248 | 0.000 | 0.993 | 28 | 1033.6 |
|  | Yichang | 2.476 | 0.000 | -0.083 | 0.000 | 2.480 | 0.000 | 0.993 | 30 | 946.2 |
|  | Jingmen | 2.558 | 0.000 | -0.079 | 0.000 | 1.630 | 0.019 | 0.981 | 32 | 929.5 |
|  | Xianning | 1.933 | 0.000 | -0.059 | 0.000 | 6.804 | 0.027 | 0.964 | 33 | 849.6 |
|  | Shiyan | 2.247 | 0.000 | -0.068 | 0.000 | 2.472 | 0.000 | 0.999 | 33 | 673.2 |
|  | Xiantao | 3.034 | 0.000 | -0.097 | 0.000 | 0.350 | 0.000 | 0.997 | 31 | 584.6 |
|  | Tianmen | 3.963 | 0.000 | -0.125 | 0.000 | 0.030 | 0.164 | 0.982 | 32 | 508.0 |
|  | Enshi | 1.701 | 0.000 | -0.053 | 0.000 | 3.851 | 0.000 | 0.993 | 32 | 254.6 |
|  | Qianjiang | 2.650 | 0.000 | -0.074 | 0.000 | 0.218 | 0.178 | 0.968 | 36 | 204.4 |
|  | Shennongjia | 0.618 | 0.000 | -0.026 | 0.000 | 2.809 | 0.000 | 0.897 | 24 | 10.8 |
| China | Hubei | 8.440 | 0.000 | -0.188 | 0.000 | 0.000 | 0.389 | 0.994 | 45 | 66767.0 |
|  | Guangdong | 2.988 | 0.000 | -0.091 | 0.000 | 0.800 | 0.001 | 0.993 | 33 | 1386.8 |
|  | Henan | 2.657 | 0.000 | -0.085 | 0.000 | 1.960 | 0.000 | 0.996 | 31 | 1306.9 |
|  | Zhejiang | 2.102 | 0.000 | -0.070 | 0.000 | 8.069 | 0.000 | 0.986 | 30 | 1239.5 |
|  | Hunan | 2.452 | 0.000 | -0.080 | 0.000 | 2.732 | 0.000 | 0.995 | 31 | 1049.2 |
|  | Anhui | 2.637 | 0.000 | -0.086 | 0.000 | 1.720 | 0.000 | 0.996 | 31 | 1019.6 |
|  | Jiangxi | 3.001 | 0.000 | -0.097 | 0.000 | 0.653 | 0.000 | 0.995 | 31 | 970.8 |
|  | Shandong | 1.805 | 0.000 | -0.033 | 0.000 | 3.731 | 0.024 | 0.981 | 55 | 853.7 |
|  | Jiangsu | 2.510 | 0.000 | -0.080 | 0.000 | 1.385 | 0.000 | 0.999 | 31 | 646.3 |
|  | Chongqing | 1.946 | 0.000 | -0.058 | 0.000 | 4.343 | 0.000 | 0.998 | 34 | 579.5 |
|  | Sichuan | 2.061 | 0.000 | -0.058 | 0.000 | 2.660 | 0.000 | 0.999 | 36 | 534.9 |
|  | Heilongjiang | 3.490 | 0.000 | -0.108 | 0.000 | 0.088 | 0.000 | 0.997 | 32 | 491.2 |
|  | Beijing | 2.257 | 0.000 | -0.066 | 0.000 | 1.354 | 0.000 | 0.996 | 34 | 408.5 |
|  | Shanghai | 2.320 | 0.000 | -0.073 | 0.000 | 1.144 | 0.000 | 0.994 | 32 | 344.9 |
|  | Hebei | 2.559 | 0.000 | -0.076 | 0.000 | 0.505 | 0.000 | 0.996 | 34 | 318.7 |
|  | Fujian | 1.805 | 0.000 | -0.060 | 0.000 | 3.929 | 0.000 | 0.994 | 30 | 300.8 |
|  | Guangxi | 1.773 | 0.000 | -0.053 | 0.000 | 2.984 | 0.000 | 0.998 | 33 | 252.9 |
|  | Shaanxi | 1.848 | 0.000 | -0.063 | 0.000 | 3.075 | 0.000 | 0.995 | 29 | 250.1 |
|  | Yunnan | 1.735 | 0.000 | -0.056 | 0.000 | 2.625 | 0.000 | 0.988 | 31 | 176.3 |
|  | Hainan | 2.284 | 0.000 | -0.068 | 0.000 | 0.554 | 0.000 | 0.993 | 33 | 170.9 |
|  | Guizhou | 3.632 | 0.000 | -0.118 | 0.000 | 0.023 | 0.004 | 0.994 | 31 | 152.6 |
|  | Tianjin | 2.642 | 0.000 | -0.075 | 0.000 | 0.158 | 0.000 | 0.997 | 35 | 136.6 |
|  | Shanxi | 2.260 | 0.000 | -0.075 | 0.000 | 0.598 | 0.000 | 0.994 | 30 | 136.4 |
|  | Liaoning | 1.759 | 0.000 | -0.058 | 0.000 | 1.784 | 0.000 | 0.997 | 30 | 123.9 |
|  | HongKong | 2.025 | 0.000 | -0.025 | 0.011 | 0.162 | 0.069 | 0.987 | 80 | 151.2 |
|  | Jilin | 2.978 | 0.000 | -0.099 | 0.000 | 0.074 | 0.023 | 0.983 | 30 | 95.7 |
|  | Gansu | 1.857 | 0.000 | -0.064 | 0.000 | 1.159 | 0.000 | 0.994 | 29 | 93.7 |
|  | Xinjiang | 2.583 | 0.000 | -0.078 | 0.000 | 0.123 | 0.000 | 0.993 | 33 | 77.5 |
|  | Neimenggu | 1.853 | 0.000 | -0.056 | 0.000 | 0.728 | 0.000 | 0.996 | 33 | 75.8 |
|  | Ningxia | 2.235 | 0.000 | -0.066 | 0.000 | 0.258 | 0.000 | 0.992 | 34 | 72.9 |
|  | Taiwan | NA | NA | NA | NA | NA | NA | NA | NA | NA |
|  | Qinghai | 1.163 | 0.000 | -0.050 | 0.000 | 1.560 | 0.000 | 0.947 | 23 | 19.0 |
|  | Macao | 0.963 | 0.000 | -0.037 | 0.000 | 1.184 | 0.000 | 0.946 | 26 | 10.4 |
|  | Xizang | NA | NA | NA | NA | NA | NA | NA | NA | NA |
| China |  | 7.419 | 0.000 | -0.172 | 0.000 | 0.000 | 0.199 | 0.996 | 43 | 79899.1 |

\*Starting dates: January 11<sup>th</sup> for Wuhan and Hubei; January 13<sup>th</sup> for the whole China; and January 19<sup>th</sup> for the rest of Chinese provinces. Ending dates: all datasets for China end at February 29<sup>th</sup>.

**Table 2.** Evaluate the error rates of the inflection-point estimations based on the infection-time relationship (ITR) with PLEC (power-law function with exponential cutoff) model

| Data and Regions | | Estimated Parameters | | Evaluation of $T_{max}$ | | | | | Evaluation of $I_{max}$ | | | |
| --- | --- | --- | --- | --- | --- | --- | --- | --- | --- | --- | --- | --- |
| Data | Country /Province | $T_{max}$ | $I_{max}$ | Observed ($T_{max}$) | Lower (95%) | Upper (95%) | 95% | Error Rate (%) | Observed (Mar 6) | Error Rate (%) | Observed (Mar 12) | Error Rate (%) |
| Hubei | Wuhan | 48 | 49193.4 | 48137 | 46733.7 | 51653.1 | Yes | 2.1% | 49797 | -1.2% | 49986 | -1.6% |
|  | Xiaogan | 32 | 3534.6 | 3465 | 3357.9 | 3711.3 | Yes | 2.0% | 3518 | 0.5% | 3518 | 0.5% |
|  | Huanggang | 32 | 2979.4 | 2899 | 2830.4 | 3128.4 | Yes | 2.7% | 2907 | 2.4% | 2907 | 2.4% |
|  | Jingzhou | 32 | 1607.0 | 1574 | 1526.7 | 1687.4 | Yes | 2.1% | 1580 | 1.7% | 1580 | 1.7% |
|  | Ezhou | 32 | 1413.2 | 1385 | 1342.5 | 1483.9 | Yes | 2.0% | 1394 | 1.4% | 1394 | 1.4% |
|  | Suizhou | 29 | 1336.7 | 1296 | 1269.9 | 1403.5 | Yes | 3.0% | 1307 | 2.2% | 1307 | 2.2% |
|  | Xiangyang | 27 | 1210.0 | 1170 | 1149.5 | 1270.5 | Yes | 3.3% | 1175 | 2.9% | 1175 | 2.9% |
|  | Huangshi | 28 | 1033.6 | 997 | 981.9 | 1085.3 | Yes | 3.5% | 1015 | 1.8% | 1015 | 1.8% |
|  | Yichang | 30 | 946.2 | 914 | 898.9 | 993.5 | Yes | 3.4% | 931 | 1.6% | 931 | 1.6% |
|  | Jingmen | 32 | 929.5 | 918 | 883.0 | 976.0 | Yes | 1.2% | 928 | 0.2% | 928 | 0.2% |
|  | Xianning | 33 | 849.6 | 836 | 807.1 | 892.1 | Yes | 1.6% | 836 | 1.6% | 836 | 1.6% |
|  | Shiyan | 33 | 673.2 | 671 | 639.5 | 706.9 | Yes | 0.3% | 672 | 0.2% | 672 | 0.2% |
|  | Xiantao | 31 | 584.6 | 571 | 555.4 | 613.8 | Yes | 2.3% | 575 | 1.6% | 575 | 1.6% |
|  | Tianmen | 32 | 508.0 | 495 | 482.6 | 533.4 | Yes | 2.6% | 496 | 2.4% | 496 | 2.4% |
|  | Enshi | 32 | 254.6 | 251 | 241.9 | 267.3 | Yes | 1.4% | 252 | 1.0% | 252 | 1.0% |
|  | Qianjiang | 36 | 204.4 | NA | 194.2 | 214.6 | No | NA | 198 | 3.1% | 198 | 3.1% |
|  | Shennongjia | 24 | 10.8 | 10 | 10.3 | 11.3 | No | 7.4% | 11 | -1.9% | 11 | -1.9% |
| Success Rates (%) |  |  |  |  |  |  | 88.2% | 88.2% |  | 100% |  | 100% |
| China | Hubei | 45 | 66767.0 | 65187 | 63428.7 | 70105.4 | Yes | 2.4% | 67592 | -1.2% | 67786 | -1.5% |
|  | Guangdong | 33 | 1386.8 | 1333 | 1317.5 | 1456.1 | Yes | 3.9% | 1352 | 2.5% | 1356 | 2.2% |
|  | Henan | 31 | 1306.9 | 1267 | 1241.6 | 1372.2 | Yes | 3.1% | 1272 | 2.7% | 1273 | 2.6% |
|  | Zhejiang | 30 | 1239.5 | 1175 | 1177.5 | 1301.5 | No | 5.2% | 1215 | 2.0% | 1215 | 2.0% |
|  | Hunan | 31 | 1049.2 | 1011 | 996.7 | 1101.7 | Yes | 3.6% | 1018 | 3.0% | 1018 | 3.0% |
|  | Anhui | 31 | 1019.6 | 989 | 968.6 | 1070.6 | Yes | 3.0% | 990 | 2.9% | 990 | 2.9% |
|  | Jiangxi | 31 | 970.8 | 934 | 922.3 | 1019.3 | Yes | 3.8% | 935 | 3.7% | 935 | 3.7% |
|  | Shandong | 55 | 853.7 | NA | 811.0 | 896.4 | No | NA | 758 | 11.2% | 760 | 11.0% |
|  | Jiangsu | 31 | 646.3 | 631 | 614.0 | 678.6 | Yes | 2.4% | 631 | 2.4% | 631 | 2.4% |
|  | Chongqing | 34 | 579.5 | 575 | 550.5 | 608.5 | Yes | 0.8% | 576 | 0.6% | 576 | 0.6% |
|  | Sichuan | 36 | 534.9 | 531 | 508.2 | 561.6 | Yes | 0.7% | 539 | -0.8% | 539 | -0.8% |
|  | Heilongjiang | 32 | 491.2 | 480 | 466.6 | 515.8 | Yes | 2.3% | 481 | 2.1% | 482 | 1.9% |
|  | Beijing | 34 | 408.5 | 399 | 388.1 | 428.9 | Yes | 2.3% | 422 | -3.3% | 436 | -6.7% |
|  | Shanghai | 32 | 344.9 | 334 | 327.7 | 362.1 | Yes | 3.2% | 342 | 0.8% | 346 | -0.3% |
|  | Hebei | 34 | 318.7 | 311 | 302.8 | 334.6 | Yes | 2.4% | 318 | 0.2% | 318 | 0.2% |
|  | Fujian | 30 | 300.8 | 293 | 285.8 | 315.8 | Yes | 2.6% | 296 | 1.6% | 296 | 1.6% |
|  | Guangxi | 33 | 252.9 | 251 | 240.3 | 265.5 | Yes | 0.8% | 252 | 0.4% | 252 | 0.4% |
|  | Shaanxi | 29 | 250.1 | 245 | 237.6 | 262.6 | Yes | 2.0% | 245 | 2.0% | 245 | 2.0% |
|  | Yunnan | 31 | 176.3 | 174 | 167.5 | 185.1 | Yes | 1.3% | 174 | 1.3% | 174 | 1.3% |
|  | Hainan | 33 | 170.9 | 168 | 162.4 | 179.4 | Yes | 1.7% | 168 | 1.7% | 168 | 1.7% |
|  | Guizhou | 31 | 152.6 | 146 | 145.0 | 160.2 | Yes | 4.3% | 146 | 4.3% | 146 | 4.3% |
|  | Tianjin | 35 | 136.6 | 135 | 129.8 | 143.4 | Yes | 1.2% | 136 | 0.4% | 136 | 0.4% |
|  | Shanxi | 30 | 136.4 | 132 | 129.6 | 143.2 | Yes | 3.2% | 133 | 2.5% | 133 | 2.5% |
|  | Liaoning | 30 | 123.9 | 121 | 117.7 | 130.1 | Yes | 2.3% | 125 | -0.9% | 125 | -0.9% |
|  | HongKong | 80 | 151.2 | NA | 143.6 | 158.8 | No | NA | 107 | 29.2% | 131 | 13.4% |
|  | Jilin | 30 | 95.7 | 91 | 90.9 | 100.5 | Yes | 4.9% | 93 | 2.8% | 93 | 2.8% |
|  | Gansu | 29 | 93.7 | 91 | 89.0 | 98.4 | Yes | 2.9% | 102 | -8.9% | 127<sup>*(1)</sup> | -35.5% |
|  | Xinjiang | 33 | 77.5 | 76 | 73.6 | 81.4 | Yes | 1.9% | 76 | 1.9% | 76 | 1.9% |
|  | Neimenggu | 33 | 75.8 | 75 | 72.0 | 79.6 | Yes | 1.1% | 75 | 1.1% | 75 | 1.1% |
|  | Ningxia | 34 | 72.9 | 71 | 69.3 | 76.5 | Yes | 2.6% | 75 | -2.9% | 75 | -2.9% |
|  | Taiwan | NA | NA | NA | NA | NA | NA | NA | 45 | NA | 49 | NA |
|  | Qinghai | 23 | 19.0 | 18 | 18.1 | 20.0 | No | 5.3% | 18 | 5.3% | 18 | 5.3% |
|  | Macao | 26 | 10.4 | 10 | 9.9 | 10.9 | Yes | 3.8% | 10 | 3.8% | 10 | 3.8% |
|  | Xizang | NA | NA | NA | NA | NA | NA | NA | 1 | NA | 1 | NA |
| Success Rates (%) |  |  |  |  |  |  | 85.3% | 85.3% |  | 85.3% |  | 82.4% |
| China |  | 43 | 79899.1 | 77785 | 75904.1 | 83894.1 | Yes | 2.6% | 80718 | -1.0% | 81003 | -1.4% |

\*Tibet (Xizang) had 1 case only and cannot be modeled with PLEC, and is excluded from computing the rates.

Listed in Table 1 included the PLEC parameters (*w, d, c*) and the *p*-values [*w*(*p*), *d*(*p*), *c*(*p*)] from estimating the corresponding parameters. These *p*-values measure the significance levels for estimating these parameters: a *p*-value<0.05 indicating that the goodness-of-fitting to the model is statistically satisfactory from the perspective of the particular parameter. *R*^2^ (*R*-squared) is of limited reference value for non-linear fitting problem, but it also confirmed the goodness-of-fitting as illustrated by the *p*-values for individual parameters. Obviously, the most important items in Table 1 are the *I*_max_ and corresponding *T*_max_, the *maximal infection number* (eqn. 3) and the corresponding *inflection time point* (eqn. 4), respectively.

Table 1 shows that PLEC fitted to virtually all datasets satisfactorily in terms of the *p*-values except for a few negligible parameter cases (*p*-value>0.05 in 4 out of 52). It should be noted that the 4 parameter cases that failed to pass significance tests only means that the parameter values may have too high standard error (variability) and the failure does not necessarily mean that the model structure failed. Therefore, the model could still be used for estimating *T*_max_ and *I*_max_, as long as the tests with actual observations are satisfactory, as explained below. Fig 1 shows an example of fitting the PLEC model based on the total infections in China from January 12^th^ to February 29^th^.

**Fig 1.**
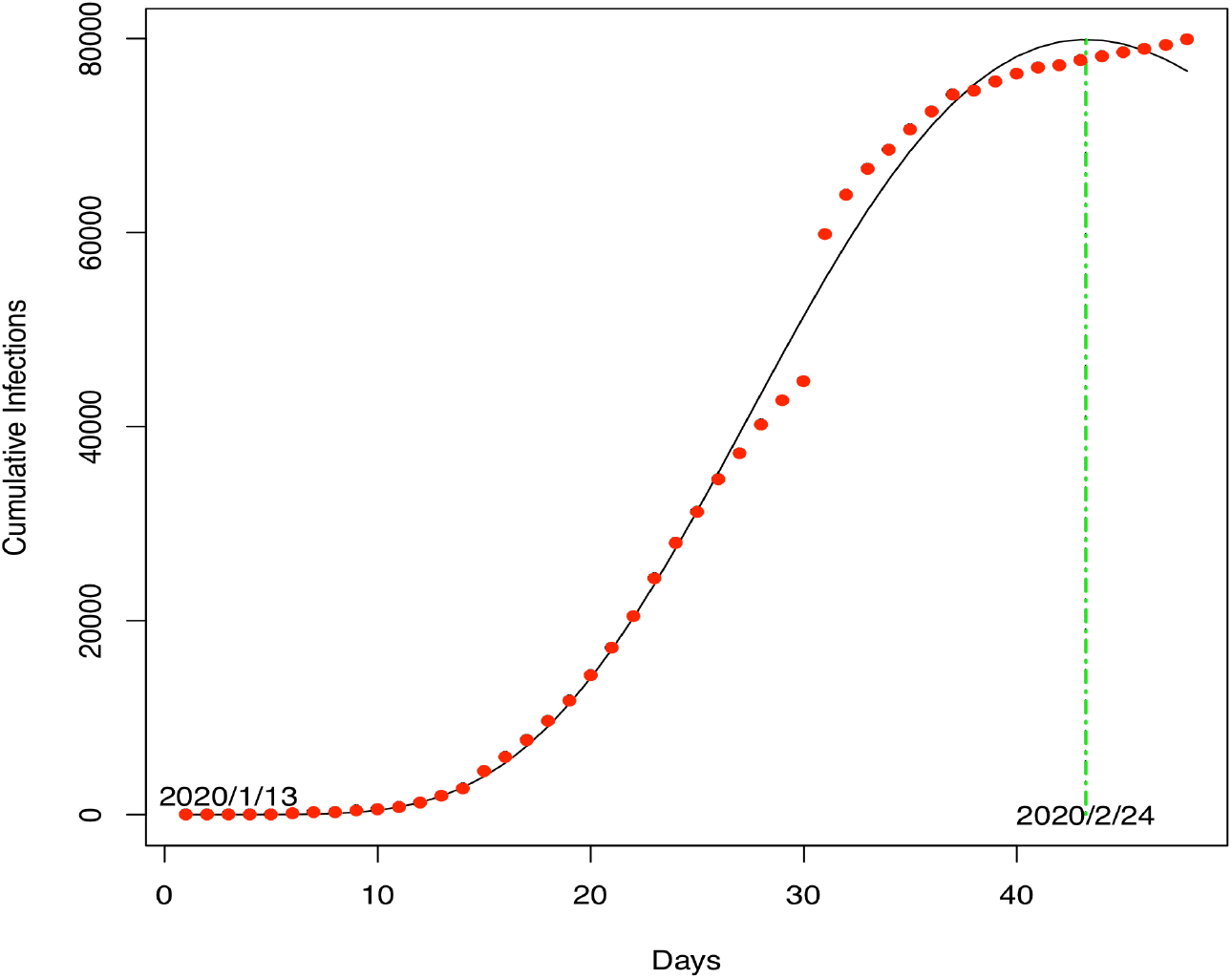
The PLEC model fitted to the COVID-19 infections in China (X-axis: Day 1=January 13^th^ 2020): the dots represent for observed infection numbers and the curve for the PLEC model.

On the whole model level, the datasets of Tibet (Xizang) and Taiwan failed to fit the PLEC model. The failure for Tibet dataset is expected since there was only one infection case and fitting a model is neither necessary nor possible; hence it was excluded from the performance evaluation. The failure to fit the infection data of Taiwan is unknown, and we instead included it in the performance evaluation as failed estimations.

Table 2 listed the results from evaluating the validity of *T*_max_ (the left side) and *I*_max_ (the right side). Regarding *T*_max_, the PLEC model correctly estimated the occurrences of inflection time points in 88% (15 out of 17 cities) in Hubei province, and 85% (29 out of 34 provinces) at the national level in China. Regarding *I*_max_, the success rates for the 17 cities in Hubei provinces were 88%, 100%, & 100% respectively, in terms of the observed infection numbers at the dates of *T*_max_, March 6^th^ and March 12^th^, respectively. Regarding *I*_max_ for the 34 provinces in China, the success rates were 85%, 85%, & 82% respectively, in terms of the observed infection numbers at the dates of *T*_max_, March 6^th^ and March 12^th^, respectively.

In summary, the test results suggest that the proposed PLEC model successfully estimated the inflection points in terms of both {*T*_max_ & *I*_max_}, *i.e*., the *inflection time points* and *maximal infection numbers*, in more than 80% tested cases (ranged between 82%-100%). Note that the date range we used to build PLEC models was between January 19 and February 29^th^, rather than the full date range available, and the datasets of the remaining dates were used for evaluating the performance of PLEC models.

Figs 2 shows both the observed and estimated maximal infection numbers {*I*_max_} for each of the 34 provinces of China, as well as the total national number (the last pair of bars). Fig 4 shows the corresponding inflection time points {*T*_max_} of each province displayed in Fig 2. Fig 3 shows the observed and estimated maximal infection numbers {*I*_max_} for each of the 17 cities of Hubei province of China, as well as the total provincial number (the last pair of bars). Fig 5 shows the corresponding inflection time points {*T*_max_} of each city.

**Fig 2.**
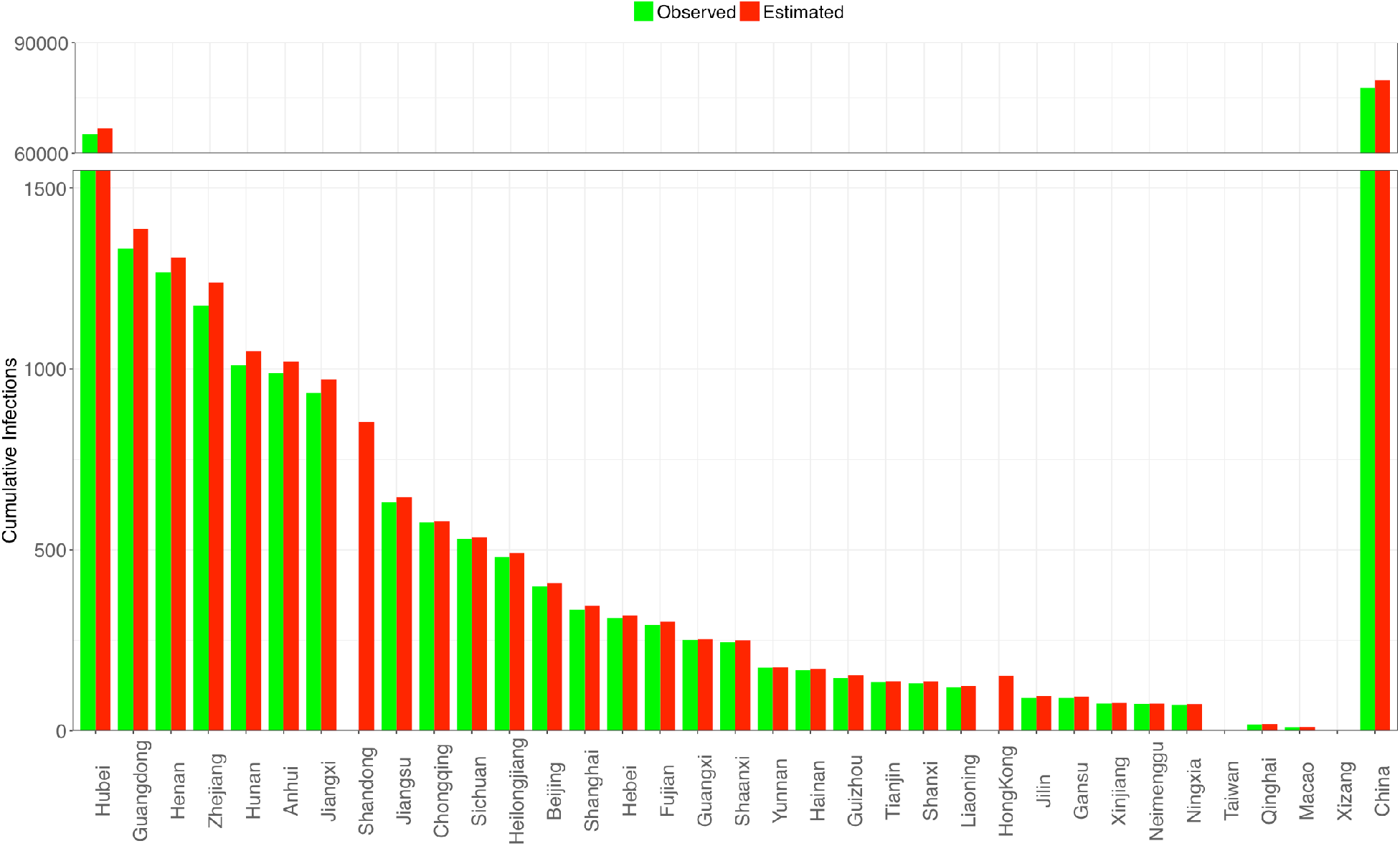
Comparisons of the observed and estimated infection time points (*I*_max_) for the 34 provinces of China; the rightmost bar represents for the inflection time point (*I*_max_) of the whole China

**Fig 3.**
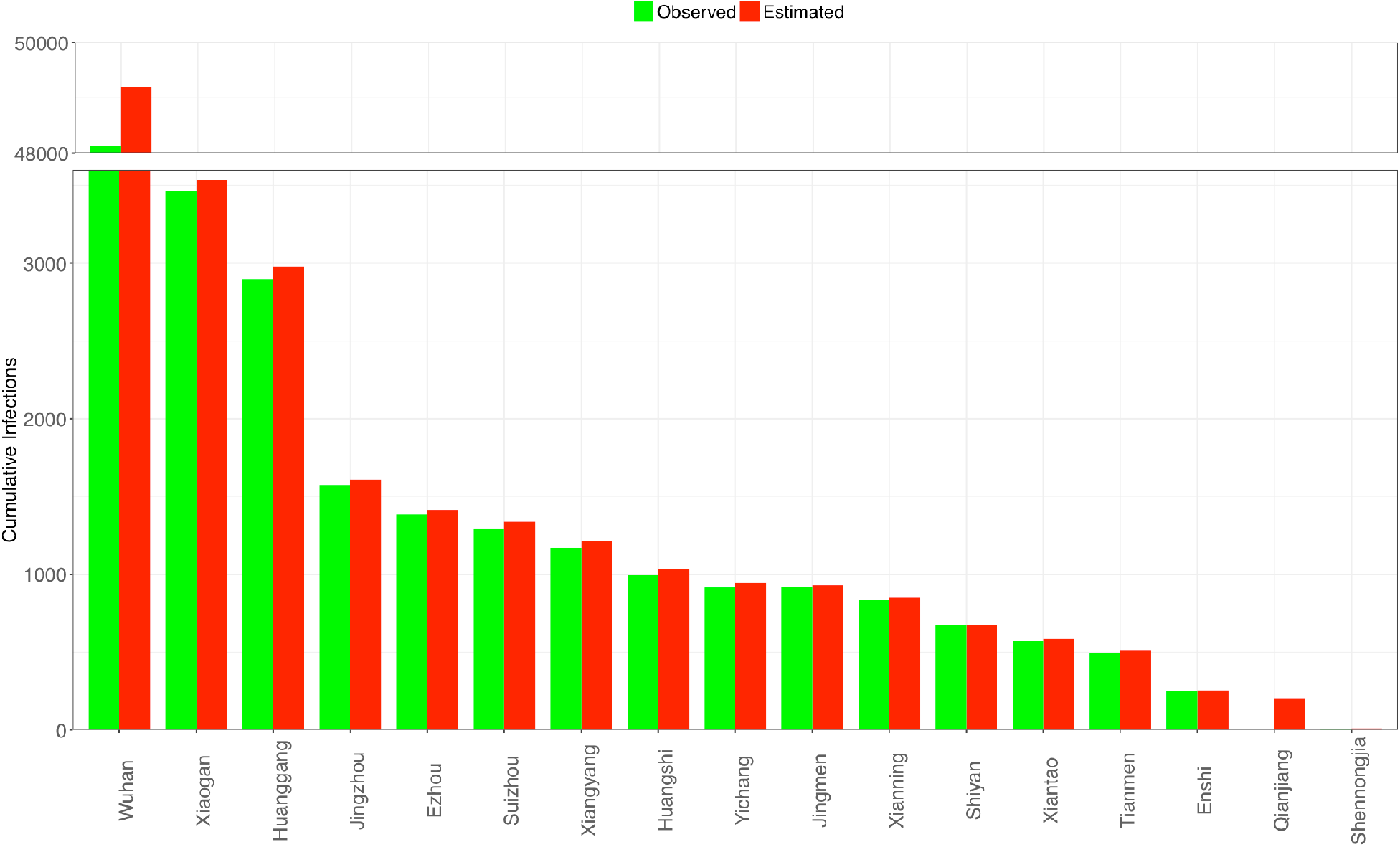
Comparisons of the observed and estimated infection time points (*I*_max_) for the 17 cities of Hubei Province, China.

**Fig 4.**
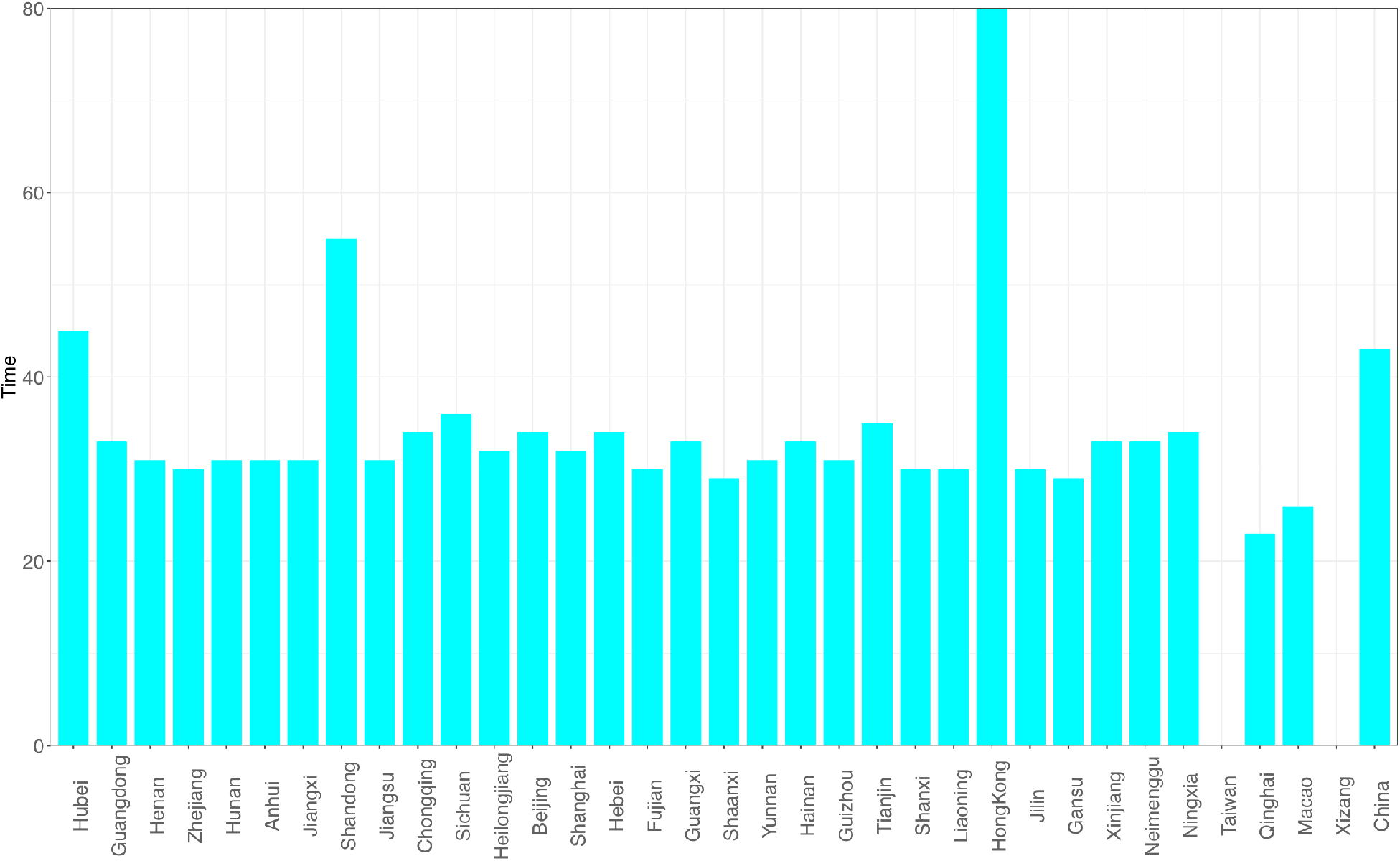
The estimated inflection time point (*T*_max_) of COVID-19 infections for 34 provinces of China; the rightmost bar represents for the inflection time point (*T*_max_) of the whole China.

**Fig 5.**
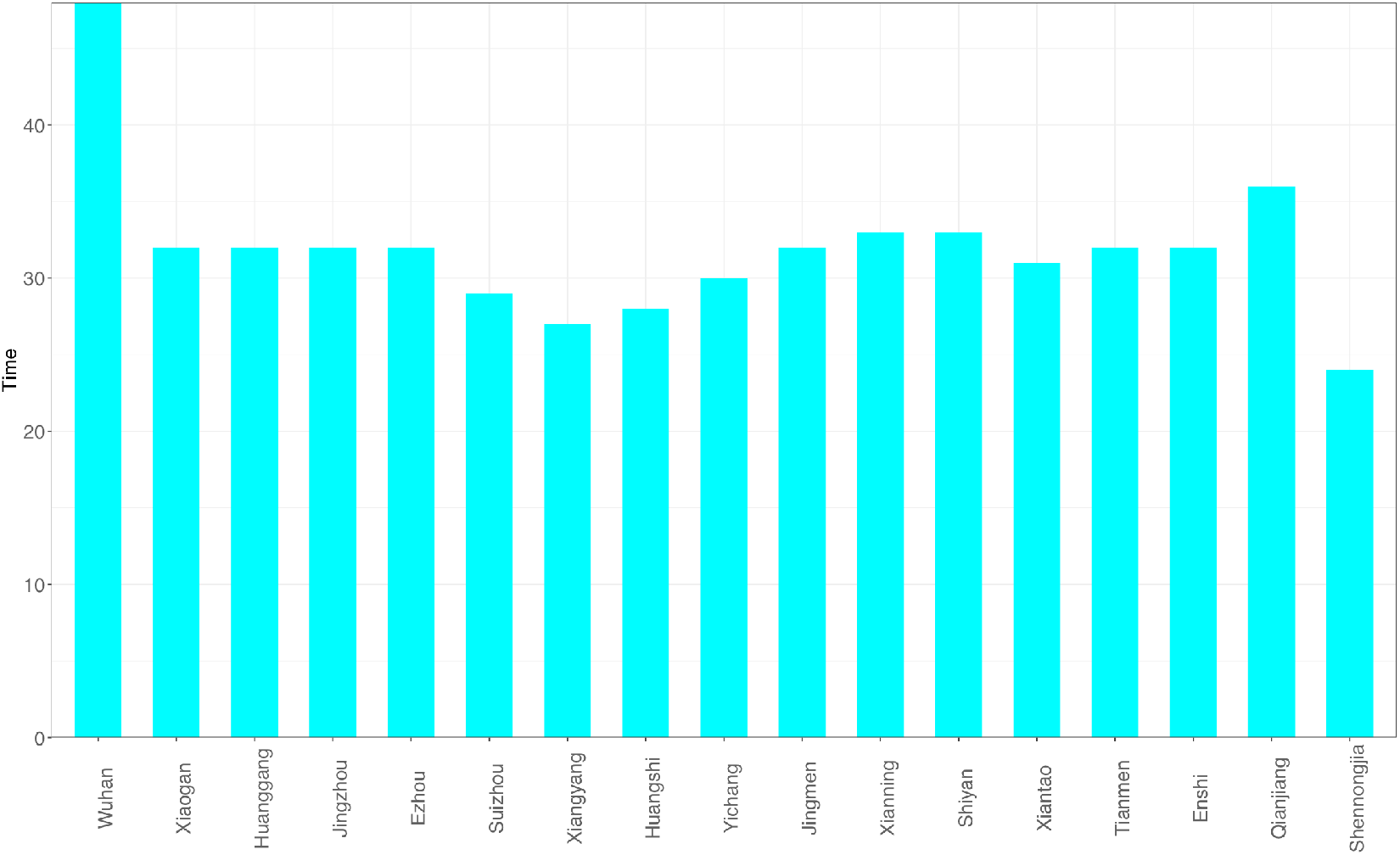
Estimated inflection time point (*T*_max_) of COVID-19 infections for the 17 cities of Hubei Province, China

## Discussion

In this section, by analyzing the failures of PLEC case by case, we try to uncover the model’s limitations. We first examine the failure exposed in previous Tables 1 & 2.

In Tables 1 & 2, the PLEC model for Shandong province performed badly with error rates about 11% was apparently due to the sudden discovery of approximately 200 cases in a local prison on a single day (Feb 20), which was approximately 1/5 of the total number in the province. The model for Gansu province also performed badly with error rates of 36%, apparently due to 33 inputs from the overseas between March 4^th^ and 8^th^, approximately 1/3 of the total number in the province. These individual cases suggest that the PLEC modeling can be sensitive to certain disruptive increases, in particular, to the increases that occupy disproportionally large proportion of the total numbers. This may be considered as the *first* limitation of the PLEC model. On the positive side, excluding the inaccuracies from these disruptive increases as well as the individual cases of Taiwan and Tibet, the PLEC model actually performed exceptionally well, reaching success rates mostly exceeding 95%.

We also fitted the PLEC model to the infection datasets of at least 14 countries (top 14 in terms of the infection numbers until March 15^th^) as well as the worldwide total infection number, and total worldwide infection number excluding China. Overall, in a majority of the cases, the PLEC model failed to obtain satisfactory predictions for the inflection points measured in {*T*_max_, *I*_max_}. To save page space, portions of the modeling results were supplied in Fig S1 of the OSI (Online Supplementary Information). From those failures, we conjecture that the failures are most likely due to the reality that in most of those countries (regions), the infections are still in the exponential growth stages. This, of course, suggests another possible limitation of the PLEC model, namely, the PLEC model may require a sufficiently long period of data points to successfully reveal the inflection points. Like all mathematical models, PLEC model is no silver bullet. Without seeing a sign of inflection point, it may not be possible for the PLEC model to reveal its existence.

Still a third limitation of the PLEC model is that it may only detect a single inflection point for a dataset. In other words, it is only applicable for detecting the inflection point for unimodal curve, as explained in the material and methods section. In the case of coronaviruses, this may not be an issue given that the SARS outbreak was also unimodal. In case of multiple-peak recurrence or seasonal outbreaks, it should be possible to divide multi-modal curve into multiple unimodal curves. Therefore, this limitation should be much less serious than it appears.

Given the above-discussed limitations of the PLEC model, we also applied the tipping point detection technique based on Fisher information (Sundstrom *et al*. 2017) for determining the inflection points, but failed to detecting any inflection (tipping) points in the datasets of COVID-19 infections. As shown in Table S1, virtually all Fisher Information (FI) values at every time window of the time series of COVID-19 equal 4, which indicates no existence of tipping points. This additional exploratory analysis not only revealed the difficulty of the problem for detecting the inflection points, but also suggested the value of PLEC model given that it successfully estimated the inflection points in terms of both {*T*_max_ & *I*_max_} on the city, provincial and national *scales* in China. To some extent, the failures of PLEC model at large scales of country should mostly due to the limitation of currently available datasets, rather than the model *per se*.

Finally, we would like to mention one potentially very important point regarding the prediction of inflection points, but is beyond the scope of this study, that is, the potentially critical influences of public-health intervention/prevention measures on the occurrences of inflection points. Such a study may only be possible after the COVID-19 worldwide pandemic is over in our opinion.

## Data Availability

All data analyzed are available in public domain and the source links are provided in the manuscript.

## Acknowledgements and Disclaims

I am indebted to the data collection and computational support from Lianwei Li and Wendy Li of the Computational Biology and Medical Ecology Lab, the Chinese Academy of Sciences. I am responsible for all the interpretations of the results, including possible errors. However, I am not responsible for any direct or indirect inferences from this article, including those from partial quotes of the text, figures and/or tables.

## Online Supplementary Information (OSI) for

Ma (2020) A Simple Mathematical Model for Estimating the Inflection Points of COVID-19 Outbreaks

**Fig S1.**
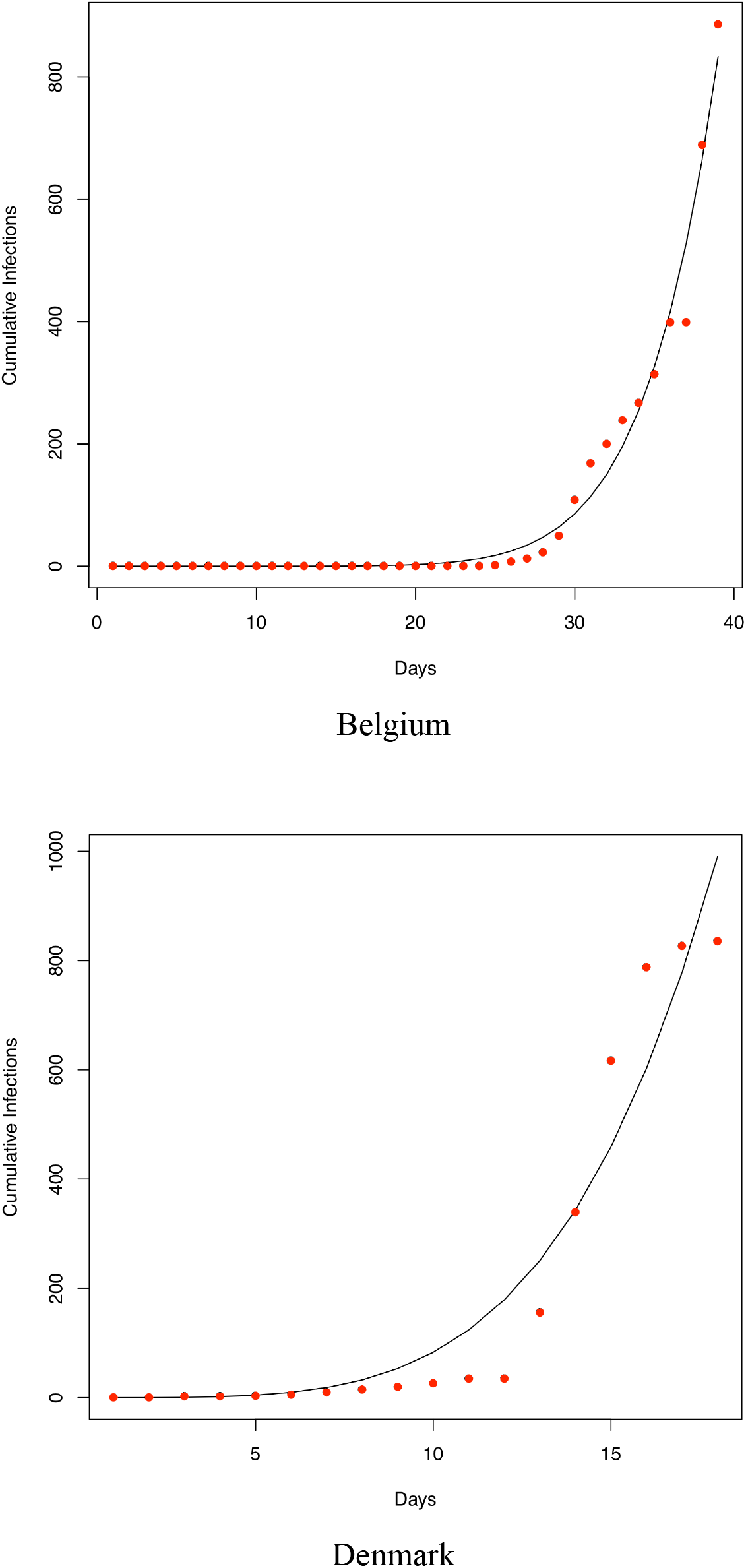
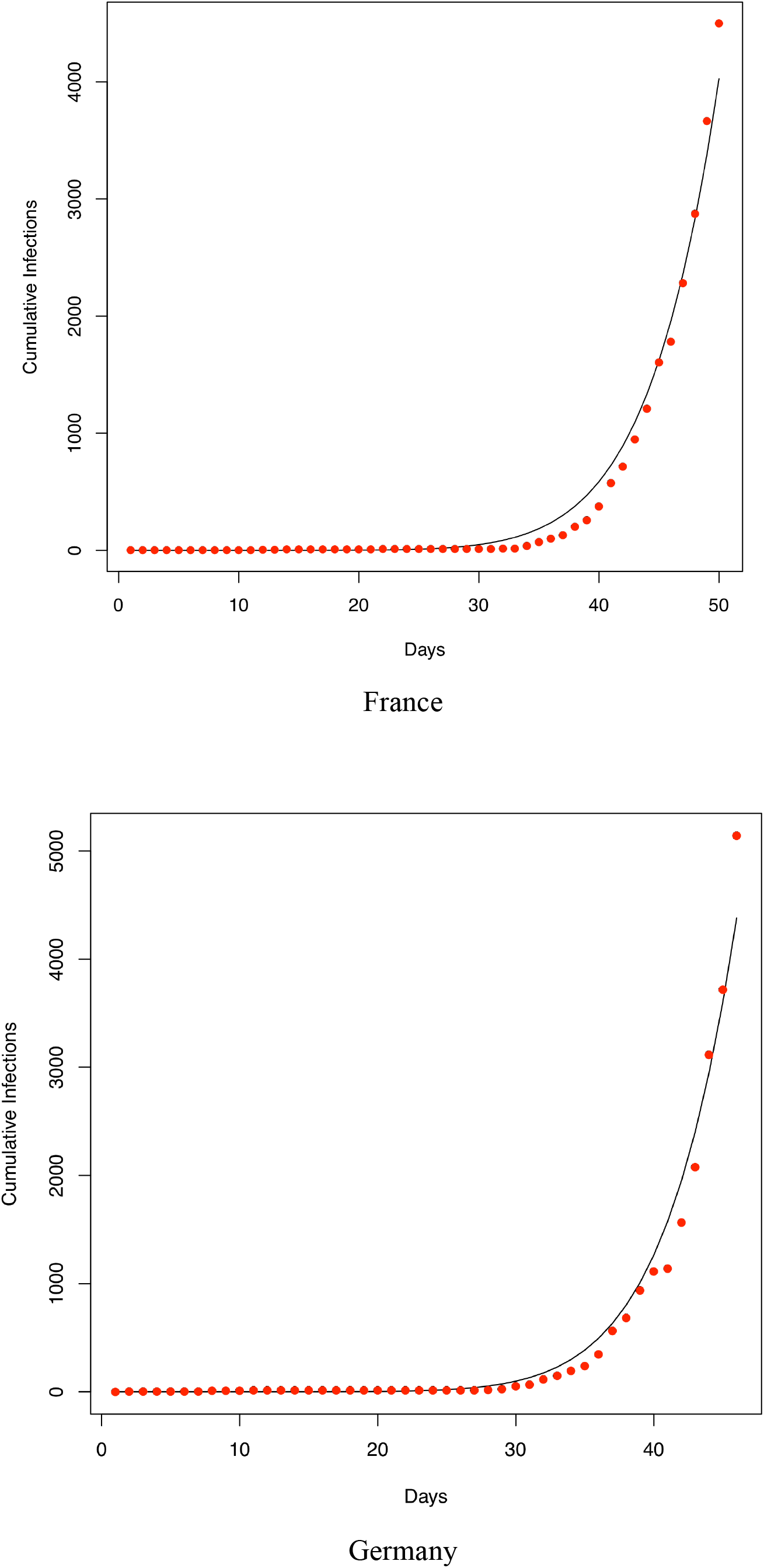
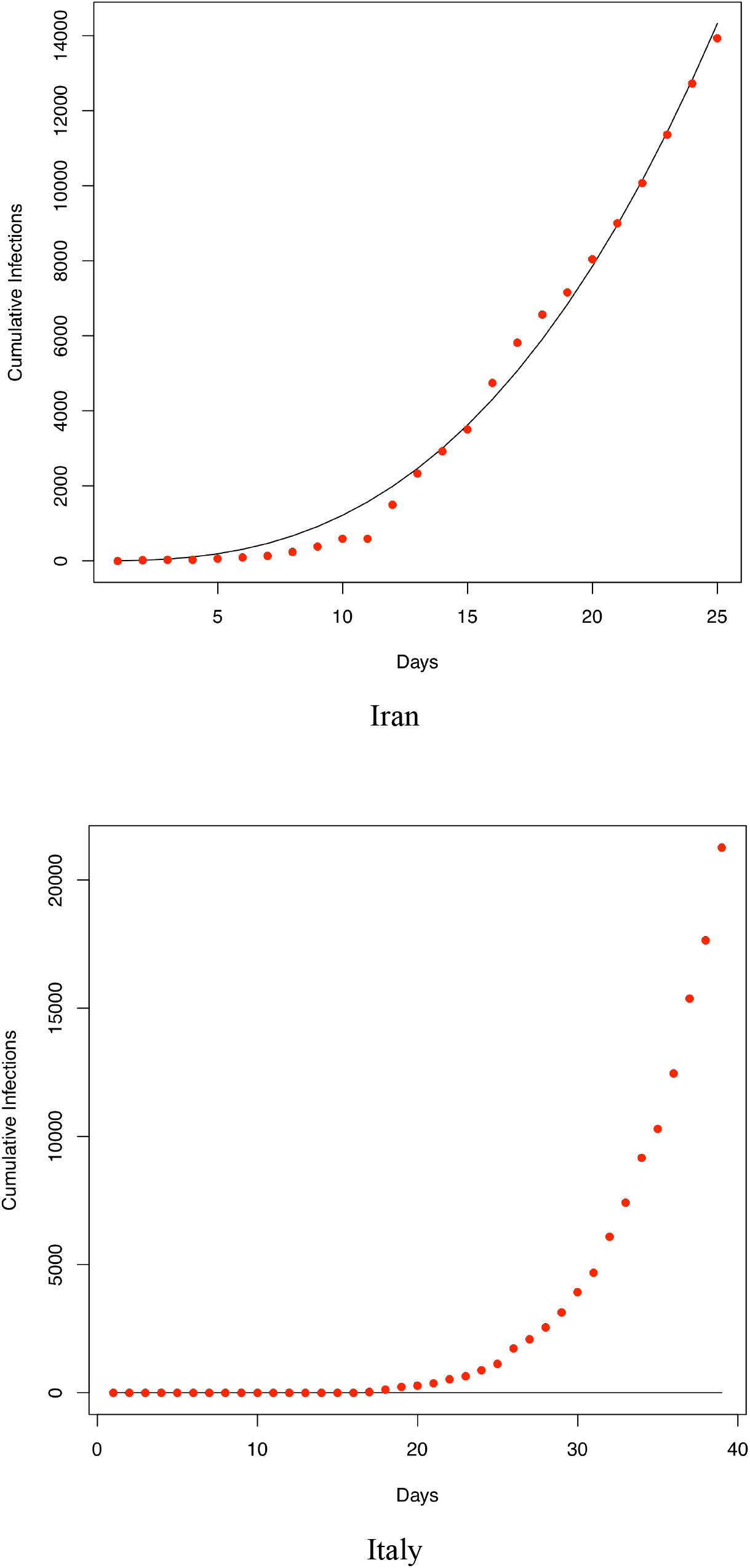
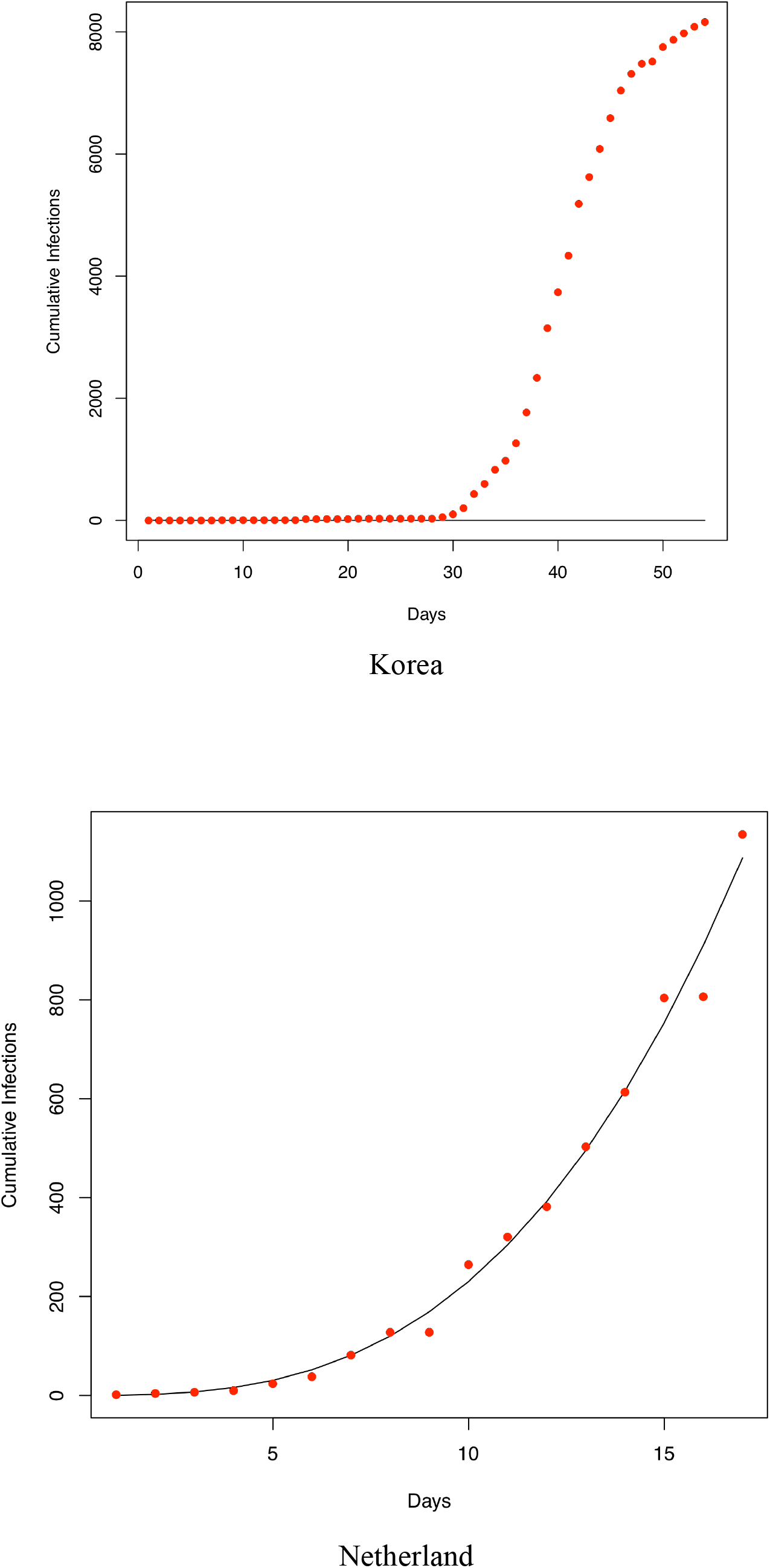
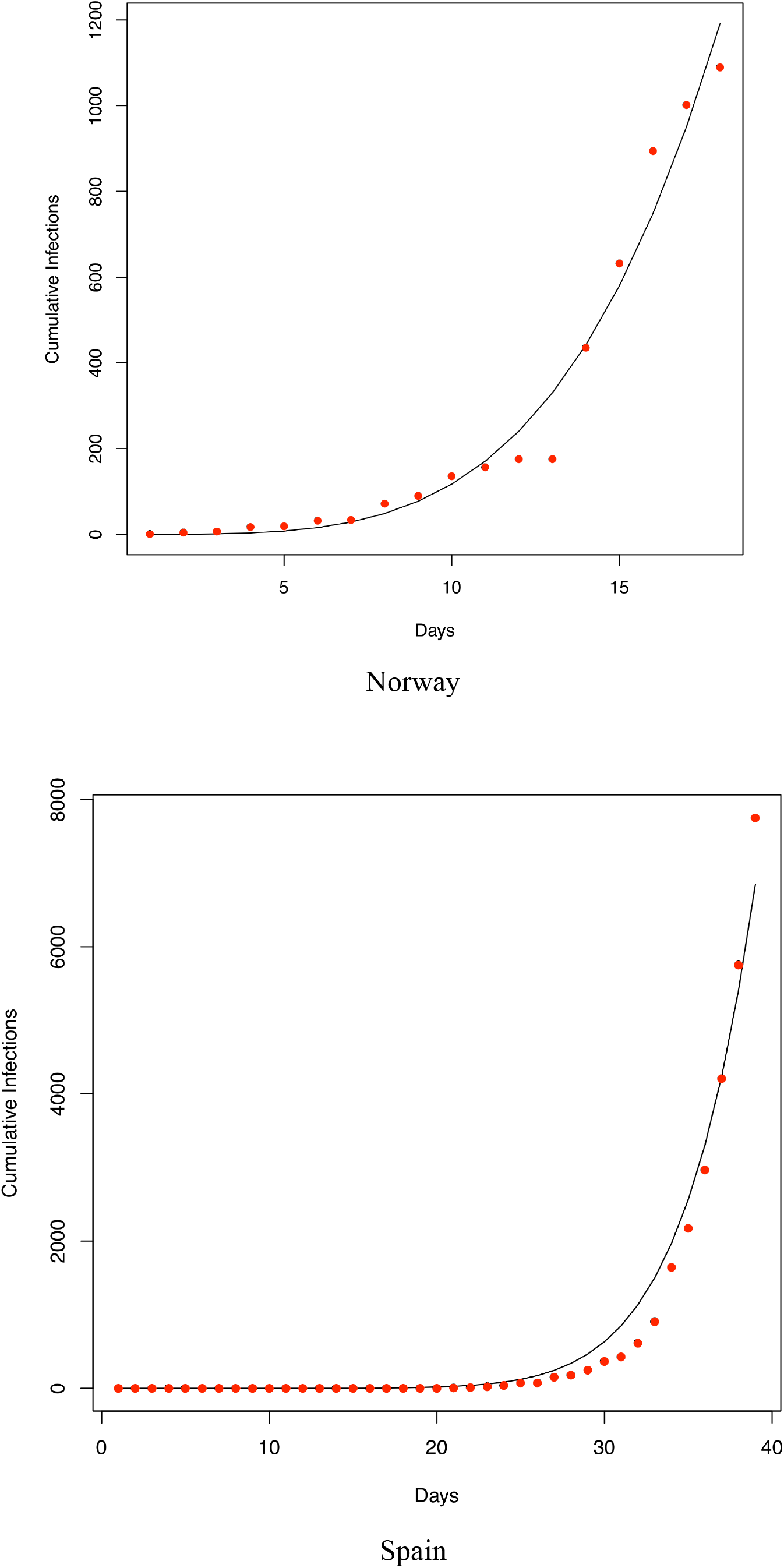
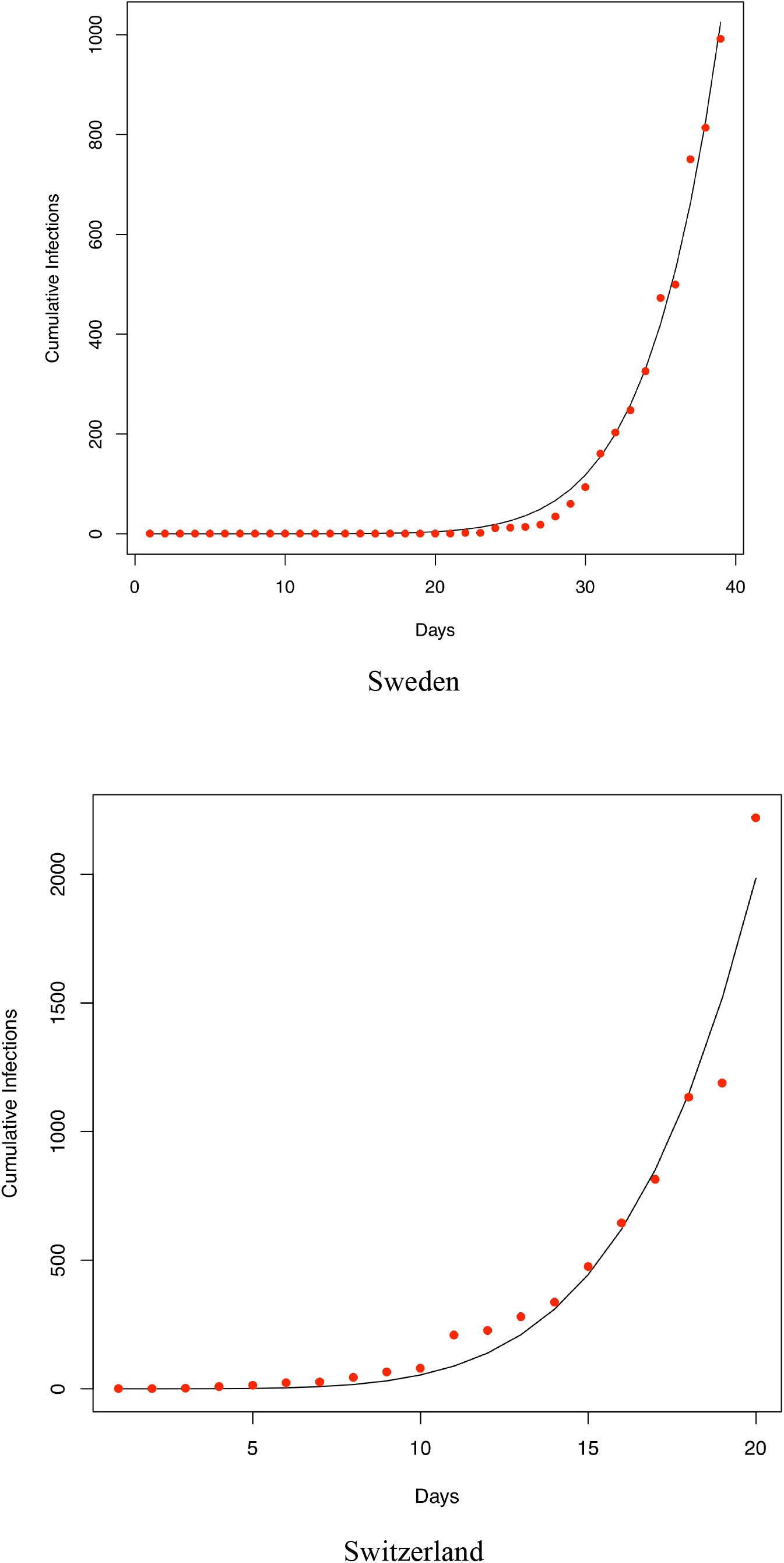
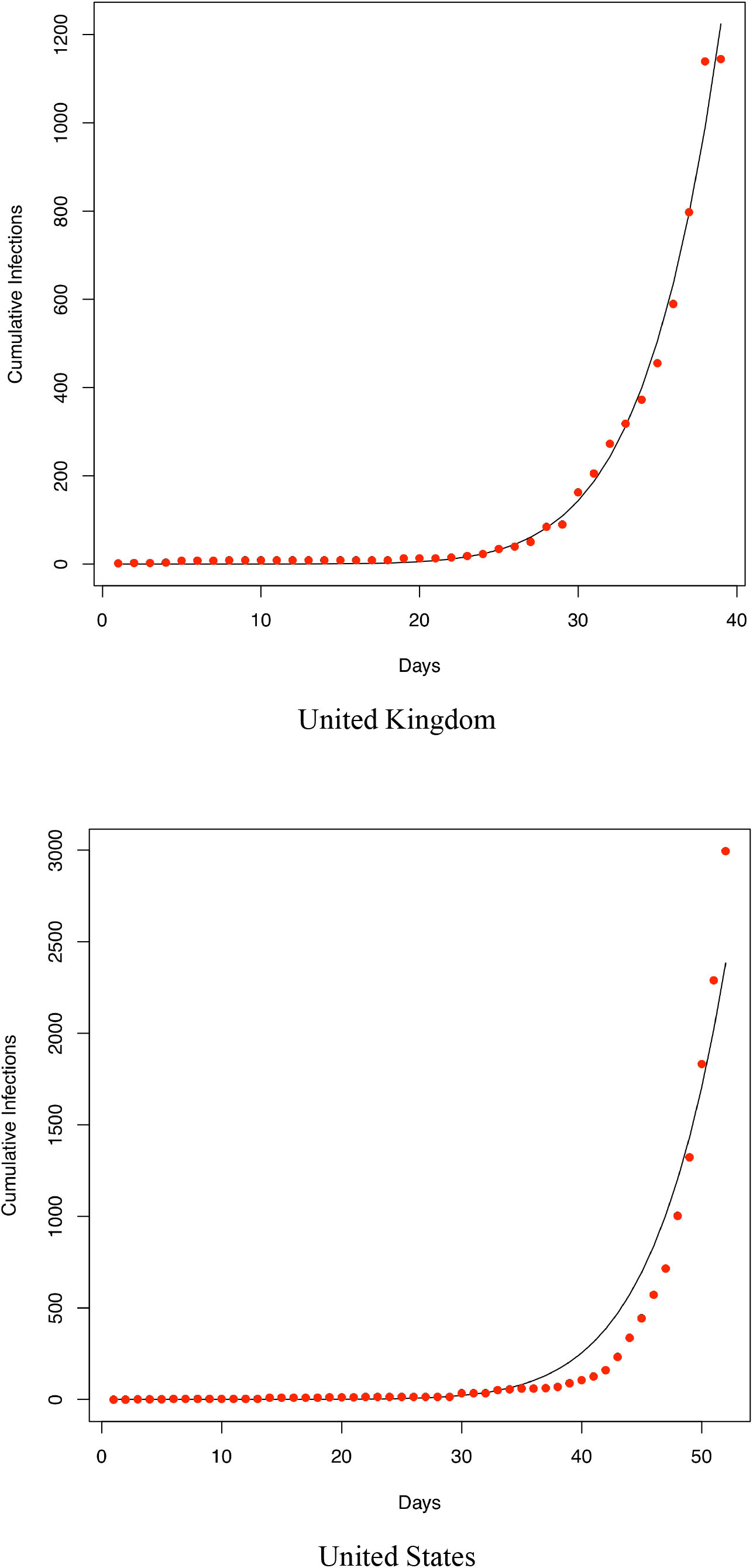
Fitting the PLEC (power law with exponential cutoff) model to the COVID-19 infections of 14 countries worldwide (which were selected based on the number of infections until Feb 15^th^). The datasets were collected between January 20^th^ and March 15^th^)

**Table 1.** The Fisher Information (FI) metric for detecting tipping points: computed for each time windows [based on the COVID-19 infection datasets of Wuhan, Hubei (between Jan 11 to Feb 29), China and World (between Jan 13 to Feb 29), and the rest of the provinces (between Jan 19 to Feb 29)]

| Data | Country /Province | 1 | 2 | 3 | 4 | 5 | ..... | 23 | 24 | 25 |
| --- | --- | --- | --- | --- | --- | --- | --- | --- | --- | --- |
| Hubei | Wuhan | 4 | 4 | 4 | 4 | 4 | ..... | 4 | 4 | 4 |
|  | Xiaogan | 4 | 4 | 4 | 4 | 4 | ..... | 4 | 4 | 4 |
|  | Huanggang | 4 | 4 | 4 | 4 | 4 | ..... | 4 | 4 | 4 |
|  | Jingzhou | 4 | 4 | 4 | 4 | 4 | ..... | 4 | 4 | 4 |
|  | Ezhou | 4 | 4 | 4 | 4 | 4 | ..... | 4 | 4 | 4 |
|  | Suizhou | 4 | 4 | 4 | 4 | 4 | ..... | 4 | 4 | 4 |
|  | Xiangyang | 4 | 4 | 4 | 4 | 4 | ..... | 4 | 4 | 4 |
|  | Huangshi | 4 | 4 | 4 | 4 | 4 | ..... | 4 | 4 | 4 |
|  | Yichang | 4 | 4 | 4 | 4 | 4 | ..... | 4 | 4 | 4 |
|  | Jingmen | 4 | 4 | 4 | 4 | 4 | ..... | 4 | 4 | 4 |
|  | Xianning | 4 | 4 | 4 | 4 | 4 | ..... | 4 | 4 | 4 |
|  | Shiyan | 4 | 4 | 4 | 4 | 4 | ..... | 4 | 4 | 4 |
|  | Xiantao | 4 | 4 | 4 | 4 | 4 | ..... | 4 | 4 | 4 |
|  | Tianmen | 4 | 4 | 4 | 4 | 4 | ..... | 4 | 4 | 4 |
|  | Enshi | 4 | 4 | 4 | 4 | 4 | ..... | 4 | 4 | 4 |
|  | Qianjiang | 4 | 4 | 4 | 4 | 4 | ..... | 4 | 4 | 4 |
|  | Shennongjia | 4 | 4 | 4 | 4 | 4 | ..... | 4 | 4 | 4 |
| China | Hubei | 4 | 4 | 4 | 4 | 4 | ..... | 4 | 4 | 4 |
|  | Guangdong | 4 | 4 | 4 | 4 | 4 | ..... | 4 | 4 | 4 |
|  | Henan | 4 | 4 | 4 | 4 | 4 | ..... | 4 | 4 | 4 |
|  | Zhejiang | 4 | 4 | 4 | 4 | 4 | ..... | 4 | 4 | 4 |
|  | Hunan | 4 | 4 | 4 | 4 | 4 | ..... | 4 | 4 | 4 |
|  | Anhui | 4 | 4 | 4 | 4 | 4 | ..... | 4 | 4 | 4 |
|  | Jiangxi | 4 | 4 | 4 | 4 | 4 | ..... | 4 | 4 | 4 |
|  | Shandong | 4 | 4 | 4 | 4 | 4 | ..... | 4 | 4 | 4 |
|  | Jiangsu | 4 | 4 | 4 | 4 | 4 | ..... | 4 | 4 | 4 |
|  | Chongqing | 4 | 4 | 4 | 4 | 4 | ..... | 4 | 4 | 4 |
|  | Sichuan | 4 | 4 | 4 | 4 | 4 | ..... | 4 | 4 | 4 |
|  | Heilongjiang | 4 | 4 | 4 | 4 | 4 | ..... | 4 | 4 | 4 |
|  | Beijing | 4 | 4 | 4 | 4 | 4 | ..... | 4 | 4 | 4 |
|  | Shanghai | 4 | 4 | 4 | 4 | 4 | ..... | 4 | 4 | 4 |
|  | Hebei | 4 | 4 | 4 | 4 | 4 | ..... | 4 | 4 | 4 |
|  | Fujian | 4 | 4 | 4 | 4 | 4 | ..... | 4 | 4 | 4 |
|  | Guangxi | 4 | 4 | 4 | 4 | 4 | ..... | 4 | 4 | 4 |
|  | Shaanxi | 4 | 4 | 4 | 4 | 4 | ..... | 4 | 4 | 4 |
|  | Yunnan | 4 | 4 | 4 | 4 | 4 | ..... | 4 | 4 | 4 |
|  | Hainan | 4 | 4 | 4 | 4 | 4 | ..... | 4 | 4 | 4 |
|  | Guizhou | 4 | 4 | 4 | 4 | 4 | ..... | 4 | 4 | 4 |
|  | Tianjin | 4 | 4 | 4 | 4 | 4 | ..... | 4 | 4 | 4 |
|  | Shanxi | 4 | 4 | 4 | 4 | 4 | ..... | 4 | 4 | 4 |
|  | Liaoning | 4 | 4 | 4 | 4 | 4 | ..... | 4 | 4 | 4 |
|  | HongKong | 4 | 4 | 4 | 4 | 4 | ..... | 4 | 4 | 4 |
|  | Jilin | 4 | 4 | 4 | 4 | 4 | ..... | 4 | 4 | 4 |
|  | Gansu | 4 | 4 | 4 | 4 | 4 | ..... | 4 | 4 | 4 |
|  | Xinjiang | 4 | 4 | 4 | 4 | 4 | ..... | 4 | 4 | 4 |
|  | Neimenggu | 4 | 4 | 4 | 4 | 4 | ..... | 4 | 4 | 4 |
|  | Ningxia | 4 | 4 | 4 | 4 | 4 | ..... | 4 | 4 | 4 |
|  | Taiwan | 4 | 4 | 4 | 4 | 4 | ..... | 4 | 4 | 4 |
|  | Qinghai | 4 | 4 | 4 | 4 | 4 | ..... | 4 | 4 | 4 |
|  | Macao | 4 | 4 | 4 | 4 | 4 | ..... | 4 | 4 | 4 |
|  | Xizang | NA | NA | NA | NA | NA | NA | NA | NA | NA |
| China |  | 4 | 4 | 4 | 4 | 4 | ..... | 4 | 4 | 4 |
| World |  | 4 | 4 | 4 | 4 | 4 | ..... | 4 | 4 | 4 |

